# Predictors and Risk Score for Immune Checkpoint-Inhibitor-Associated Myocarditis Severity

**DOI:** 10.1101/2024.06.02.24308336

**Authors:** John R. Power, Charles Dolladille, Benay Ozbay, Adrien Procureur, Stephane Ederhy, Nicolas L. Palaskas, Lorenz H. Lehmann, Jennifer Cautela, Pierre-Yves Courand, Salim S. Hayek, Han Zhu, Vlad G. Zaha, Richard K. Cheng, Joachim Alexandre, François Roubille, Lauren A. Baldassarre, Yen-Chou Chen, Alan H. Baik, Michal Laufer-Perl, Yuichi Tamura, Aarti Asnani, Sanjeev Francis, Elizabeth M. Gaughan, Peter P. Rainer, Guillaume Bailly, Danette Flint, Dimitri Arangalage, Eve Cariou, Roberta Florido, Anna Narezkina, Yan Liu, Shahneen Sandhu, Darryl Leong, Nahema Issa, Nicolas Piriou, Lucie Heinzerling, Giovanni Peretto, Shanthini M. Crusz, Nausheen Akhter, Joshua E Levenson, Isik Turker, Assié Eslami, Charlotte Fenioux, Pedro Moliner, Michel Obeid, Wei Ting Chan, Stephen M. Ewer, Seyed Ebrahim Kassaian, International ICI-Myocarditis Registry, Douglas B. Johnson, Anju Nohria, Osnat Itzhaki Ben Zadok, Javid J. Moslehi, Joe-Elie Salem

## Abstract

**Background:** Immune-checkpoint inhibitors (ICI) are associated with life-threatening myocarditis but milder presentations are increasingly recognized. The same autoimmune process that causes ICI-myocarditis can manifest concurrent generalized myositis, myasthenia-like syndrome, and respiratory muscle failure. Prognostic factors for this “cardiomyotoxicity” are lacking.

**Methods:** A multicenter registry collected data retrospectively from 17 countries between 2014-2023. A multivariable cox regression model (hazard-ratio(HR), [^95%^confidence-interval]) was used to determine risk factors for the primary composite outcome: severe arrhythmia, heart failure, respiratory muscle failure, and/or cardiomyotoxicity-related death. Covariates included demographics, comorbidities, cardio-muscular symptoms, diagnostics, and treatments. Time-dependent covariates were used and missing data were imputed. A point-based prognostic risk score was derived and externally validated.

**Results:** In 748 patients (67% male, age 23-94), 30-days incidence of the primary composite outcome, cardiomyotoxic death, and overall death were 33%, 13%, and 17% respectively. By multivariable analysis, the primary composite outcome was associated with active thymoma (HR=3.60[1.93-6.72]), presence of cardio-muscular symptoms (HR=2.60 [1.58-4.28]), low QRS-voltage on presenting electrocardiogram (HR for ≤0.5mV versus >1mV=2.08[1.31-3.30]), left ventricular ejection fraction (LVEF) <50% (HR=1.78[1.22-2.60]), and incremental troponin elevation (HR=1.86 [1.44-2.39], 2.99[1.91-4.65], 4.80[2.54-9.08], for 20, 200 and 2000-fold above upper reference limit, respectively). A prognostic risk score developed using these parameters showed good performance; 30-days primary outcome incidence increased gradually from 3.9%(risk-score=0) to 81.3%(risk-score≥4). This risk-score was externally validated in two independent French and US cohorts. This risk score was used prospectively in the external French cohort to identify low risk patients who were managed with no immunosuppression resulting in no cardio-myotoxic events.

**Conclusions:** ICI-myocarditis can manifest with high morbidity and mortality. Myocarditis severity is associated with magnitude of troponin, thymoma, low-QRS voltage, depressed LVEF, and cardio-muscular symptoms. A risk-score incorporating these features performed well.

**Trial registration number**: *NCT04294771 and NCT05454527*

**Structured Graphical Abstract:** **Key Question:** What predicts poor prognosis in ICI-associated myocarditis?

**Key Finding:** Troponin, thymoma, cardio-muscular symptoms, low QRS-voltage, and LVEF predicted negative outcomes. A risk score containing these features performed well.

**Take-Home Message:** ICI-myocarditis is part of a generalized myotoxicity and early risk-stratified early is possible.

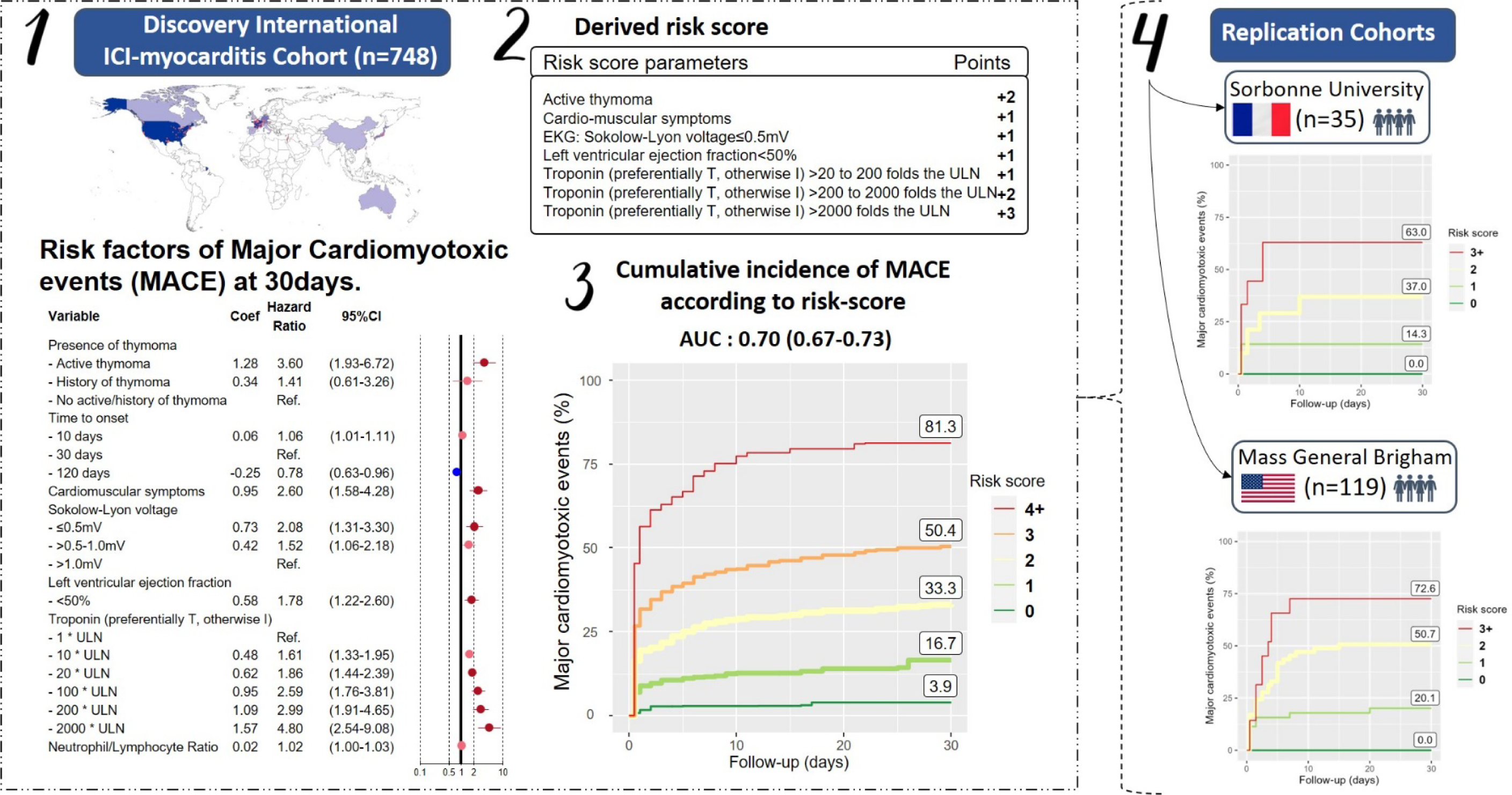

## Introduction

Immune checkpoint inhibitors (ICI) have emerged as a mainstay of cancer therapy with widespread adoption across multiple tumor types and clinical settings. Almost 50% of patients with cancer are currently eligible for ICI therapy.^1^ ICI activate the immune system with anti-tumor effects but cause immune related adverse events (irAE) in 70-90% of patients.^2–4^ IrAE can affect any organ including cardio-muscular tissues and range from mild to fatal.^1,5–8^ ICI-myocarditis, which occurs in 0.3-1% of ICI recipients, is the most fatal irAE with mortality estimated at 30-50% in early studies.^5,7,9^ However, ICI-myocarditis severity is heterogeneous; while asymptomatic cases are increasingly recognized, others patients experience life-threatening arrhythmias, cardiogenic shock, or severe concomitant myositis with respiratory muscle failure.^5,10^ Triage and management of this rare condition remains a significant challenge even for advanced multidisciplinary teams.

To date, prognostic studies of ICI-myocarditis have used pharmacovigilance datasets or cohorts less than 150 patients, without external validation and with differing results.^7,9,11,12^ Proposed predictors of severity include combination ICI treatment, elevated biomarkers, conduction delays, systolic dysfunction, inadequate immunosuppression, and simultaneous myasthenia or myositis irAE. Since these features often coexist, reconciling these data is challenging. Consequently, major knowledge gaps remain in selecting the appropriate level of care and immunosuppression strategy for ICI-myocarditis. To address this, we leveraged a web-based platform and built a multicenter registry with approximately 750 adjudicated cases of ICI-myocarditis. In this work, we use this cohort to perform a comprehensive multivariable analysis of predictors of severe outcomes in ICI-myocarditis. We then generated and externally validated a risk score to assist clinicians in risk stratification and treatment decisions.

## Methods

### Data Collection

A retrospective registry spanning 127 institutions across 17 countries containing 757 cases of ICI-myocarditis diagnosed by treating clinicians was used as a discovery cohort (Table S1, Figure S1 for collaborators). Clinical data were locally adjudicated and submitted by participating centers via a HIPPA-compliant REDCap web-based platform (IRB:181337; NCT04294771). Submissions were reviewed and cases were rejected when evidence did not meet Bonaca’s diagnostic criteria for myocarditis, a hierarchical definition incorporating the availability and strength of diagnostic data.^13^ Electrocardiograms (ECG) on presentation were independently interpreted by two cardiologists (B.O. and C.D) blinded to specific cases who performed quantitative measurements (Sokolow-Lyon Index, PR, and QRS length) and identified qualitative features (methods previously published).^14,15^ The primary outcome, major cardio-myotoxic events, was defined as any of the following: 1) severe heart failure requiring either intravenous medicines or mechanical circulatory support 2) severe arrhythmia defined as sustained ventricular arrythmia, complete heart block, sudden cardiac death, use of atropine or isoproterenol, and/or use of pacemaker/defibrillator 3) respiratory failure due to myositis-related respiratory muscles failure requiring ventilation and/or 4) death due to these conditions (henceforth referred to as cardio-myotoxicities). Respiratory muscle failure was included to reflect contemporary understanding of ICI-myocarditis as part of a generalized skeletal myositis that can impair ventilation.^5,8,16,17^ The study period ended 30 days after clinical presentation with ICI-myocarditis to avoid including unrelated endpoints. Cases were excluded if the primary outcome was met more than 24 hours prior to presentation (n=9), leaving 748 cases for analysis.

Two external independent validation ICI-myocarditis cohorts (using the updated Bonaca’s criteria) were obtained from Sorbonne University (SU, Paris, France) (prospective cohort of 35 consecutive cases presenting between April 2023 and April 2024 where the risk score was used to guide treatment; *NCT05454527*)^5,18^ and Mass General Brigham (MGB, Boston, USA) (retrospective cohort of 119 consecutive cases presenting between January 2015 and June 2022, with enough available elements to compute the replication),^19^ both collected after derivation of a risk-score from the discovery cohort. The same inclusion and exclusion criteria, and outcomes definition from the discovery cohort were applied.

## Statistical Analysis

### Explanatory model

A univariate cox regression model was used to evaluate the association of clinical variables with major cardio-myotoxic events in the discovery cohort. Independent variables spanned past medical history, objective data collected during the ICI-myocarditis evaluation, and immunosuppressant use. Troponin, creatine-kinase, cardio-muscular symptoms, left ventricular ejection fraction(LVEF), and immunosuppressant use were treated as time-dependent covariates to prevent survivorship bias. Troponin and creatine-kinase were divided by the contributing institution’s upper limit of normal (ULN). When both cardiac troponin-T and troponin-I were collected, troponin-T was preferentially used.^18^ A multivariable explanatory model tested pre-selected variables based upon available evidence and clinical experience. Multivariable analysis was performed after multiple imputation by chained equations of missing data using 50 imputed datasets. Imputed results were merged according to Rubin’s rule.^20^

### Predictive model

Starting from an empty predictive model, a stepwise selection procedure was performed using Schwartz Criteria to select the most impactful variables. The untransformed coefficients of these variables were used to derive a risk score for predicting the primary outcome with one point assigned for a coefficient of 0.5-1, two points for a coefficient of >1-1.5, and three points for a coefficient >1.5. Internal validation was performed with K-fold cross-validation with k-set of 10. External validation was performed on the two external validation ICI-myocarditis cohorts. For both internal and external validation, Harrell’s c-index assessed the risk score’s goodness-of-fit using serial models on days 1-3. Confidence intervals were obtained with bootstrap, using up to 1000 bootstrap replicates. Sensitivity analysis was performed with an unimputed dataset as well as a competing risk model to account for death from other causes. Analyses were conducted with R-v.4.2.2 for Windows, and the R-packages survival, mice, boot and rms. A two-sided p-value<0.05 was deemed significant.

## Results

### Patient Characteristics and Outcomes

Among 748 cases of ICI-myocarditis, median(interquartile-range) age was 69(61-77) and 33% were female (Table-1 for the baseline characteristics). Median time from first ICI dose to myocarditis presentation was 40(23-77) days with 21% patients receiving combination ICI and 79% receiving ICI monotherapy (usually anti-PD-1). The most common primary cancers were skin (31%), lung (24%), and genitourinary (22%); 3% had a thymoma. Half the cohort was classified as definite myocarditis, and one quarter each as probable and possible. Among those who underwent cardiac MRI(n=459), studies were consistent with myocarditis in 64% patients and when endomyocardial biopsy or autopsy was performed (n=290), histopathology was consistent with myocarditis in 67% patients. Patients without supporting MRI or histopathology met myocarditis criteria by other diagnostics; 84% had cardio-muscular symptoms within 72h of presentation, 95% had elevated troponin, 94% had abnormal ECG, 42% had depressed LVEF, and 43% had either concomitant myositis or myasthenia gravis-like syndrome (Figure-1A). Corticosteroids were given to 83% of patients and 36% received non-steroidal immunomodulators.

**Figure 1:**
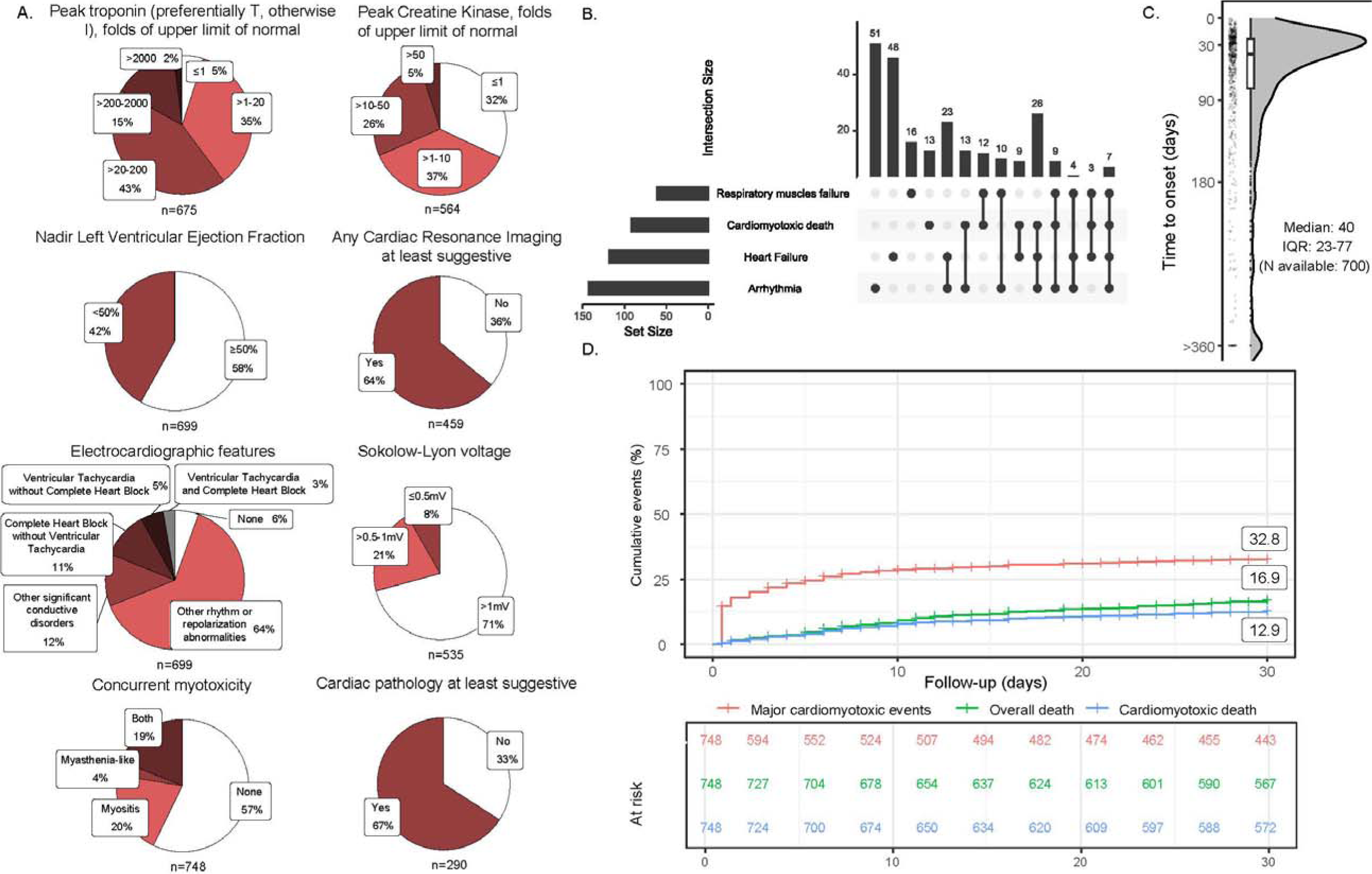
Cardiac workup and clinical event rate. Results of diagnostics used in myocarditis workup (A), major cardio-myotoxic events overlap (B), time from first ICI dose to onset of myocarditis (C), and event-curve of first major cardio-myotoxic event at 30-days (D).

**Table 1:**
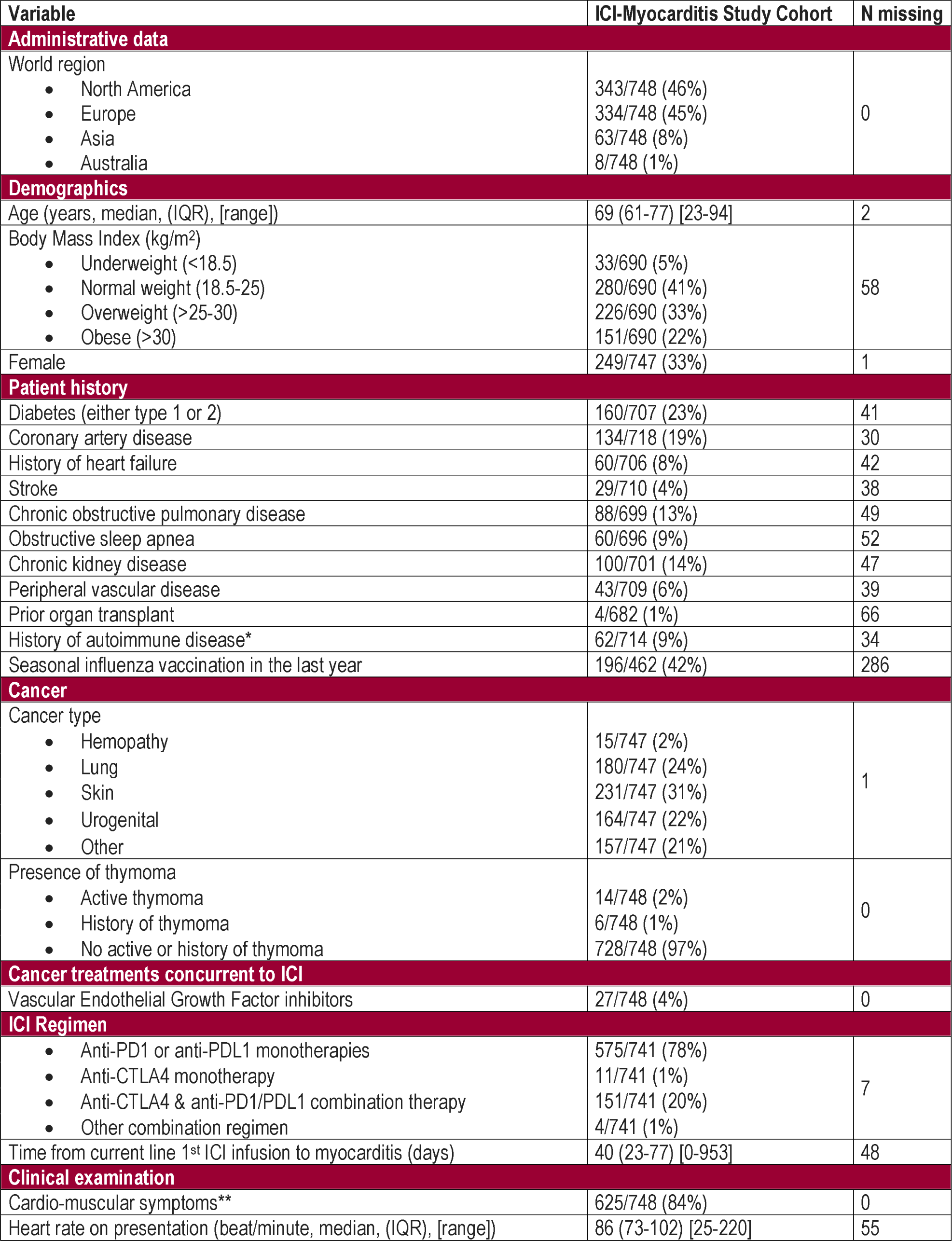
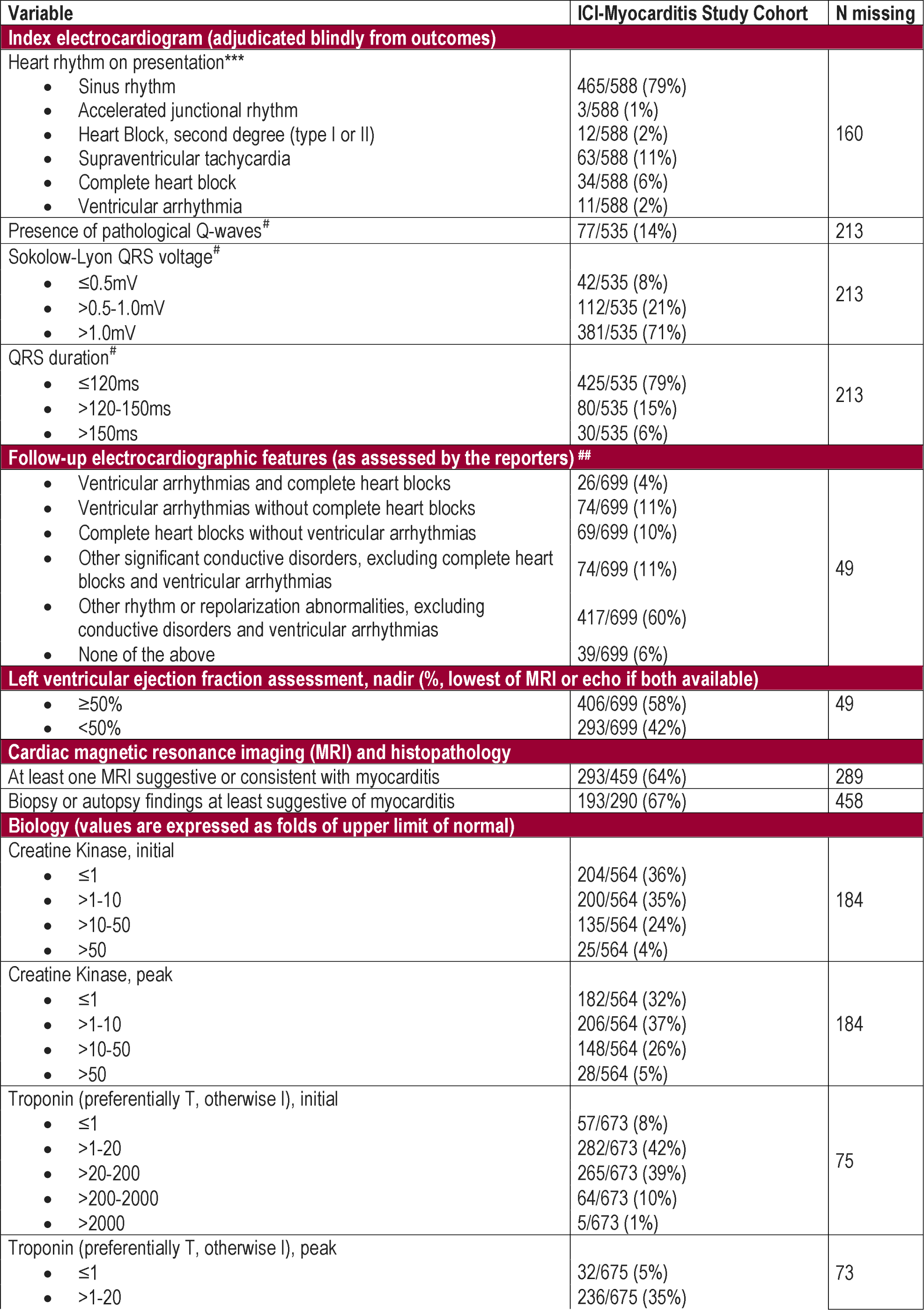
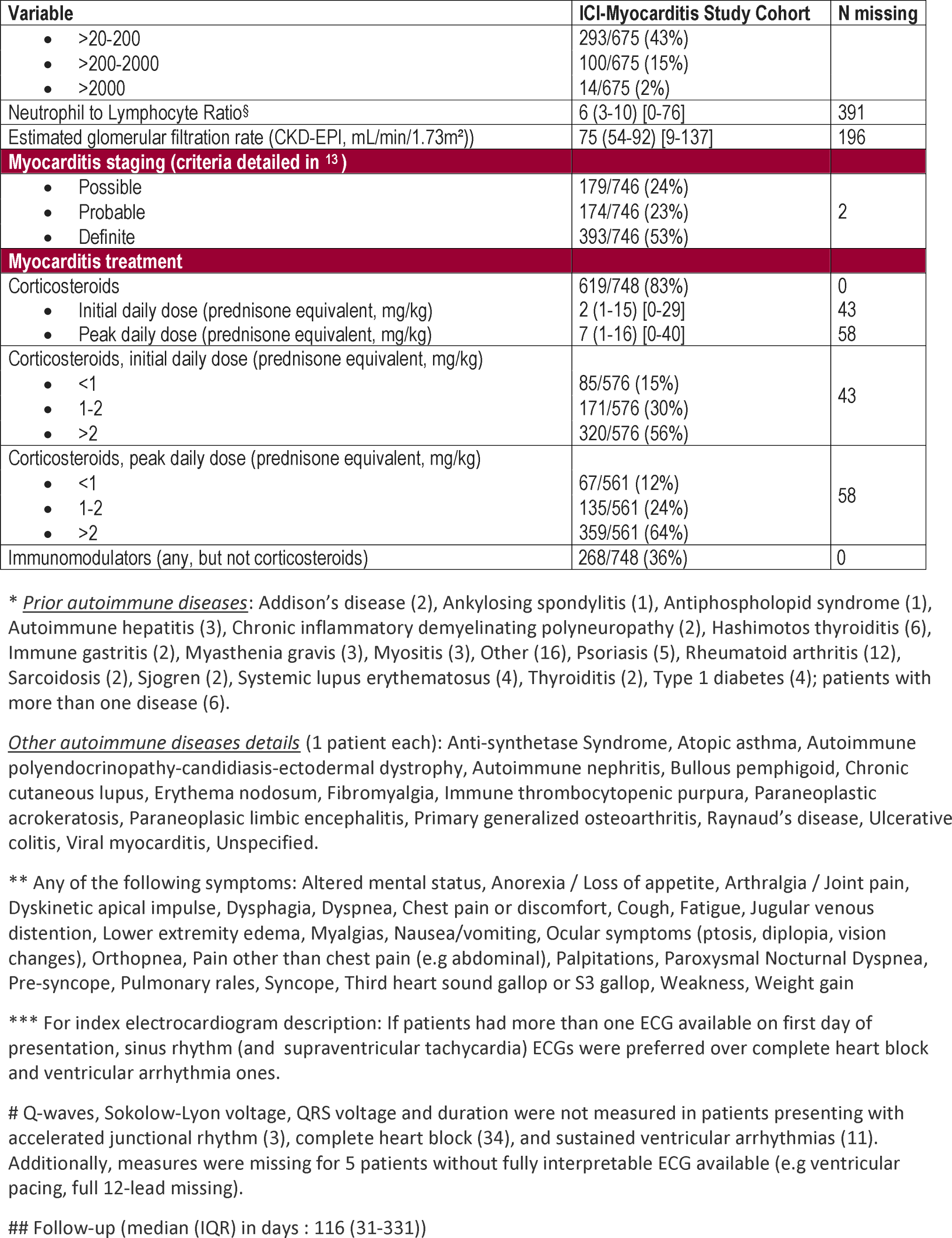
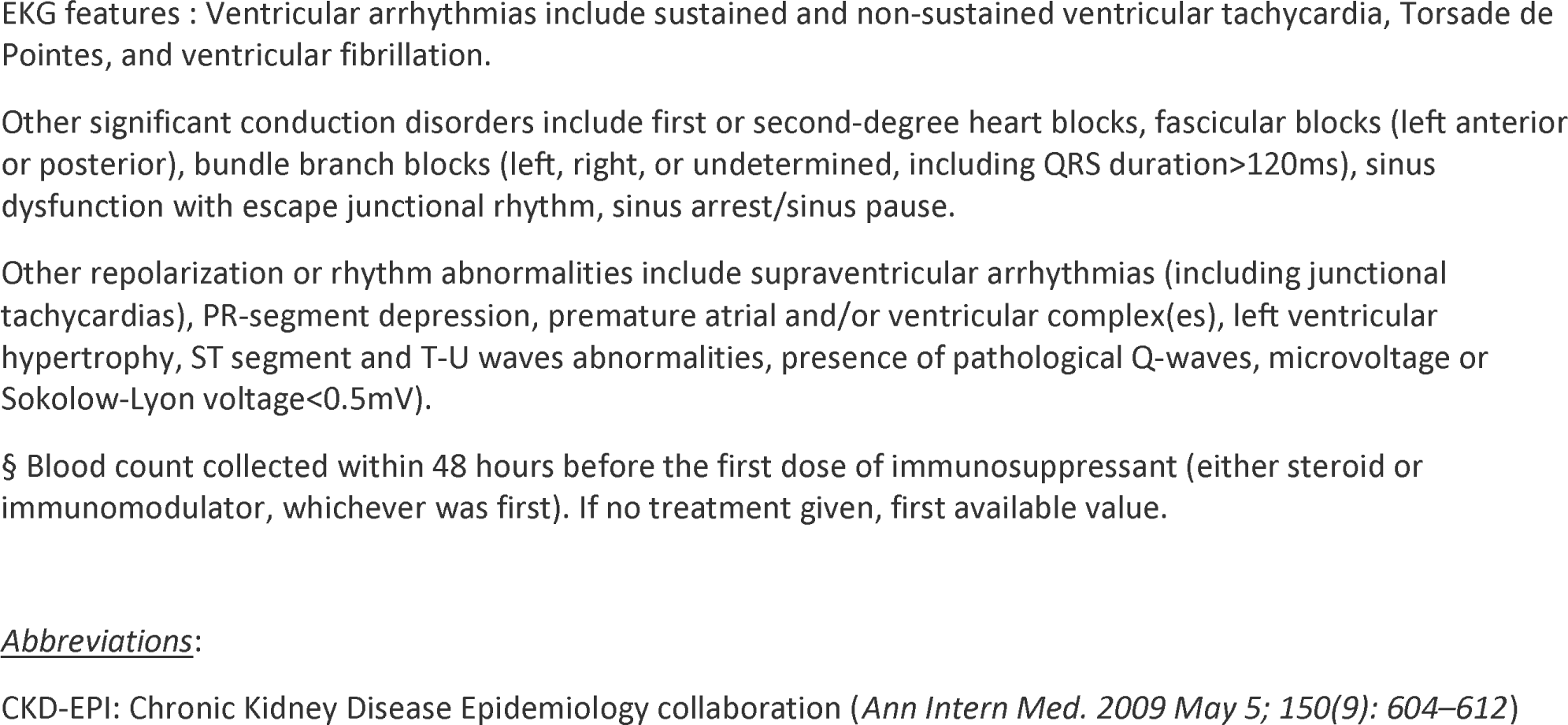
Discovery cohort characteristics (n=748)

The primary composite outcome of major cardio-myotoxic events occurred in one third of patients within one month of presentation. This included 118/748(16%) patients with heart failure, 143/748(19%) with arrhythmia, 61/748(8%) with respiratory muscle failure, and 92/748(13%) with death due to cardio-myotoxicity (Figure-1B for the intersection of primary outcome components). Most cardio-myotoxicity events and related deaths occurred early in the 30-days study period while overall death (17%) was more evenly distributed (Figure-1D for event curves).

### Explanatory Analysis

Multivariable modeling found that major cardio-myotoxic events were associated with active thymoma (HR=3.75[1.82-7.69]), recent ICI initiation (HR for 10 versus 30 days=1.06[1.01-1.11]), presenting ECG with low QRS-voltage (HR for ≤0.5mV versus >1mV =1.88[1.15-3.07]), LVEF<50% (HR=1.78[1.19-2.67]), presence of cardio-muscular symptoms (HR=2.60[1.57-4.31]) and troponin elevation (HR=1.83[1.38-2.43], 2.91 [1.76-4.81], 4.63[2.25-9.53] for 20, 200 and 2000-fold increase over upper reference limit (Figure-2A). In this model, notable variables that showed no association with severe cardio-myotoxicity included age, sex, combination ICI therapy, corticosteroid dose, and use of non-steroidal immunomodulators.

**Figure 2:**
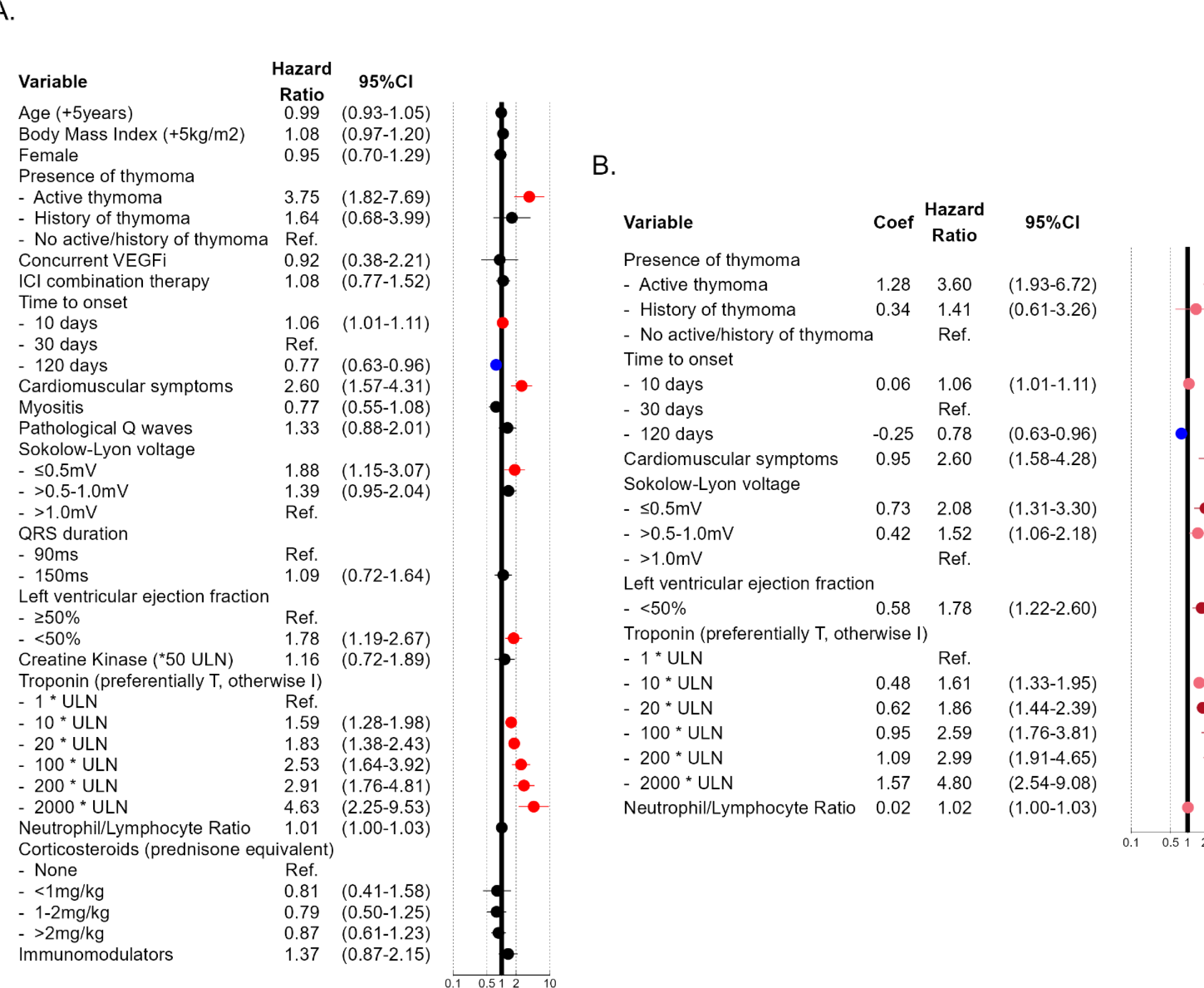
Multivariable analysis of association with 30-days major cardio-myotoxic events. Explanatory multivariable model for association with major cardio-myotoxic event (A). Predictive multivariable model for association with major cardio-myotoxic event using a stepwise selection procedure (B). *<u>Abbreviations</u>*: CI : confidence interval, coef: coefficient, ICI: immune checkpoint inhibitor, ms: millisecond, mV: millivolt, Ref: reference, ULN: upper limit of normal, VEGFi: Vascular Endothelial Growth Factor Inhibitor, Immunomodulator: non-steroidal immunomodulators only

Univariate analysis associated major cardio-myotoxic events with several features, some of which did not meet significance upon multivariable analysis. Those included high body mass index (HR=1.10[1.00-1.21]), cardio-muscular symptoms (HR=3.44[2.10-5.63]), pathological Q-waves (HR=1.96[1.34-2.89]), magnitude of QRS prolongation (HR=1.64[1.18-2.27] for 150ms vs. 90ms), elevated creatine kinase (HR for 50-fold increase over ULN=2.44[1.77-3.35]), elevated neutrophil-to-lymphocyte ratio (HR=1.03[1.01-1.04]), and treatment with non-steroidal immunomodulators (HR=1.98[1.34-2.94]) (Figure S2A). The incremental association of severe cardio-myotoxicity with continuous variables on day of presentation including biomarkers, ECG, LVEF, and time to onset are displayed in Figure S3. Similar results were found when multivariable modeling was performed on an unimputed complete dataset (Figure S2B). A competing risk analysis correcting for the effect of death due to other causes had overall unchanged findings and effect size compared to the original multivariable model (Figure S4).

### Predictive Analysis and Risk Score

A stepwise variable selection procedure identified the strongest predictors of the composite major cardio-myotoxic event outcome to be active thymoma (HR=3.60[1.93-6.72]), presence of cardio-muscular symptoms (HR=2.60[1.58-4.28]), low QRS-voltage on presenting electrocardiogram (HR for ≤0.5mV versus >1mV=2.08[1.31-3.30]), LVEF<50% (HR=1.78[1.22-2.60]), and elevated troponin (HR=1.86[1.44-2.39], 2.99[1.91-4.65], 4.80[2.54-9.08], for 20, 200 and 2000-fold increase over institution’s upper limit respectively) (Figure-2B). A risk score ranging from 0 to 8 was derived by assigning each predictor a value proportional to its respective regression coefficient: +2 for active thymoma, +1 for cardio-muscular symptoms, +1 for low QRS-voltage, +1 for LVEF<50%, and +1-3 points for degree of troponin elevation (Figure-3A). Classification of the cohort according to risk score on day of presentation showed 9% with 0 points, 29% with 1 point, 36% with 2 points, 18% with 3 points, and 4% with 4 or more points (Figure-3B). 30-days cumulative incidence of major cardio-myotoxic events was low at 3.9% for patients with risk score of 0 and increased proportionally as risk score increased with 16.7% incidence for 1 point, 33.3% for 2 points, 50.4% for 3 points, and 81.3% for 4 or more points (Figure-3C). There was a 1.6% cumulative incidence of cardio-myotoxicity related death for a risk score of 0 compared to 34.8% for a score≥4. All components of the primary outcome increased incrementally with higher risk scores (Figure-3D).

**Figure 3:**
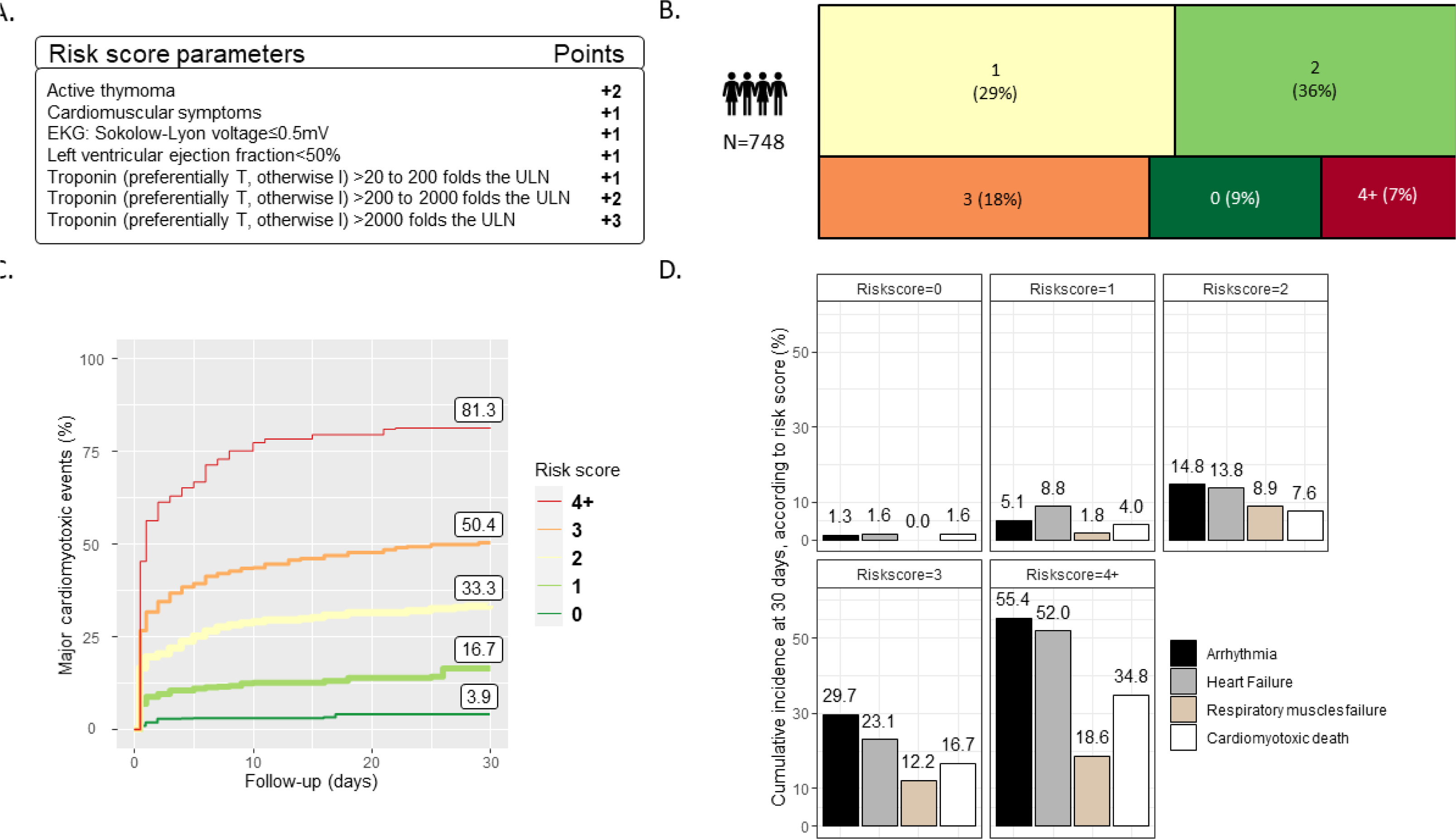
Risk score for major cardio-myotoxic events. Components of the risk score of major cardio-myotoxic events at 30-days (A). Distribution of patients according to risk score (B). Cumulative incidence of major cardio-myotoxic events according to for risk score level (C). Cumulative incidence of components of the primary composite outcome of cardio-myotoxic events (D).

### Risk Score Validation

Internal validation of the risk score showed good performance with a c-index of 0.70[0.67-0.73] on day one and remained unchanged at 0.71[0.66-0.75] by day three (Figure-S5). A score of 0 had a negative predictive value of 97%[90-100] for severe myotoxicity and 98%[91-100] for myotoxicity-related death (Table S3 for predictive properties per risk score value).

The external validation cohorts’ characteristics are detailed in Table-S2. Compared to the discovery cohort, the SU cohort had a higher proportion of score 0 (20%), but scores 1, 2, and 3+ were similar (31%, 31%, and 17%, respectively) to the discovery cohort (Figure-S6). The MGB cohort had almost no patients with score 0(<1%), but more with score 1(37%) and score 2(45%), and fewer with score 3+(18%). For risk score 0, 30-days cumulative incidence of major cardio-myotoxic events was 0% in both cohorts and increased incrementally for score 1(22.1% SU vs 20.1% MGB), score 2(39.8% SU vs 50.7% MGB), and were highest for risk score 3+(63% SU vs 72.6% MGB) (Figure-S6). In the MGB cohort, the risk score had a c-index of 0.63(0.55-0.71) on day 1 which increased to 0.72(0.63-0.80) on day 2 and 0.75(0.65-0.82) on day 3. In the SU cohort, the risk score had c-index of 0.75(0.53-0.86) on day 1 and 0.74(0.52-0.87) on day 2. In both cohorts, a risk score of 0 had 100% negative predictive value (Table-S3, Figure-S5). In the prospective SU validation cohort, all seven patients with risk score of 0 had ICI stopped but no immunosuppression given per a recently published therapeutic protocol.^5^ There was no death or major cardiomyotoxic events in these seven patients over one month.

## Discussion

ICI-myocarditis manifests with a wide range of severity. This study used a contemporary multicenter registry to follow 748 patients with ICI-myocarditis for one month after presentation, observing 13% mortality from cardio-myotoxicity. ICI-myocarditis was highly morbid with a primary composite outcome of severe heart failure, arrhythmia, respiratory muscle failure, and cardio-myotoxicity-related mortality occurring in 33% of patients. Multivariable analysis showed this primary outcome was associated with elevated troponin, thymoma, cardio-muscular symptoms, LVEF<50%, and ECG with low QRS-voltage. To avoid over-fitting and favor generalization, a simplified risk score was generated from these features which predicted adverse outcomes with good accuracy and very high negative predictive value. The risk score was externally validated in both a prospective cohort of 35 patients at Sorbonne University (Paris, France) and a retrospective cohort of 119 patients at Mass General Brigham (Boston, USA). In the prospective cohort, all patients with a risk score of zero had ICI withheld and were managed without immunosuppressants with excellent outcomes.

While early reports of ICI-myocarditis estimated a mortality of 50%, this study’s lower event rate is consistent with contemporary reports that include cases with less severe disease.^9,11,21,22^ To date, few cohort studies have analyzed ICI-myocarditis outcomes with sample sizes ranging from 33 to 147. All studies used a major adverse cardiac event (MACE) primary outcome, typically including cardiovascular death, complete heart block, ventricular arrhythmias and/or cardiogenic shock.^9,14,23–28^ Over 1-5 month follow-up, 23-64% MACE was observed, similar to our discovery cohort’s major cardiomyotoxic event rate of 33% and our validation cohorts’ rates of 31% (SU) and 41% (MGB). Our study used a 30-days surveillance period which reduced the cumulative event rate but prevented misattribution of late endpoints in this multimorbid population.

With 748 patients, this is the largest cohort of ICI-myocarditis ever studied. Unlike previous studies, this cohort also included patients with mild or sub-clinical ICI-myocarditis (∼20% of the cohort) reflecting contemporary understanding that milder asymptomatic cases exist.^5,21^ In contrast to previous studies only evaluating a limited number of predictors, our study’s size allowed for a large multivariable analysis including variables implicated in previous studies as well as novel predictors. Consequently, previous associations with severe outcomes such as combination ICI therapy or corticosteroid use, were not found to be significant.^9,24,26,27^ These findings reframe combination ICI therapy as a risk factor for developing myocarditis with little prognostic value when other variables are considered.^11,29,30^ The lack of benefit with corticosteroids is provocative given their widespread use. In the discovery cohort, it is likely that immunosuppression was prescribed to sicker patients at higher risk for adverse events. On the other hand, the SU prospective validation cohort followed a pre-defined therapeutic algorithm where patients with risk score of 0 were managed without any immunosuppressants and had no major cardiomyotoxic events at one month. It is worth considering these findings in light of ongoing debates over appropriate corticosteroid dosing in ICI-myocarditis, high rates of corticosteroid resistance, and recent evidence of higher mortality in severe cases when corticosteroids are used as first line therapy without non-steroidal immunosuppressants.^5,31,32^

The strongest predictors of adverse outcomes identified in this study – troponin levels, thymoma, low QRS-voltage, LVEF and cardio-muscular symptoms – are logical from a mechanistic perspective. Troponin release is inherent to the inflammatory myocyte necrosis of myocarditis and has been demonstrated as a surrogate for disease severity in smaller studies.^9,18,24^ While univariate analysis of creatine kinase showed significant association in this and other studies, the multivariable model did not, suggesting it may be confounded by other biomarkers, namely troponin.^18,33^ While only 14/748 (2%) of patients in this study received ICI for active thymoma, these patients had nearly 4-fold higher severe cardio-myotoxicity risk after multivariable adjustment. This is consistent with increased autoimmunity seen in thymic neoplasms and fits the leading hypothesis that ICI-myocarditis is associated with thymic dysregulation and generation of auto-reactive T-cells targeting cardio-muscular antigens and unleashed by ICI.^8,34–36^ Similarly, the prognostic value of low QRS-voltage is also based in pathophysiology as immune infiltrates have been shown to directly impact myocyte electrophysiology and induce arrhythmias.^14,37^

The results of this analysis are compelling given their clinical utility. Factors such as troponin, symptoms, ECG features, LVEF and thymoma status can be promptly ascertained in ambulatory and emergency room settings. While advanced imaging or endomyocardial biopsy results have been linked to outcomes in previous studies, the inaccessibility of these resource-intensive diagnostics makes them impractical prognostic tools.^23,25,38^ Separately, this study’s identification of risk profiles complements ongoing efforts to optimize immunosuppression for the treatment of ICI-myocarditis. For example, while ruxolitinib and abatacept have been effectively used to treat severe ICI-myocarditis, asymptomatic cases without severity infra-clinical surrogates have been managed by simply stopping ICI without steroids or other immunosuppressants with good outcomes.^5^ Two prospective randomized studies are currently testing abatacept treatment in ICI-myocarditis. While one trial only includes patients with suspected mild to moderate ICI-myocarditis without ventricular arrhythmias, high-degree atrio-ventricular blocks, or cardiogenic shock (NCT05335928), the other aims to optimize abatacept dosing in definite ICI-myocarditis with these latter severe features (NCT05195645). Using predictive risk scores like ours in future studies could identify patients with the most potential for positive treatment effect.

There are multiple potential clinical applications of this risk score. A score of 0 in combination with thorough clinical assessment may identify patients who can be managed with minimal or no immunosuppression since cardio-myotoxic events are rare in this group. Since over 14% of patients with a risk score of 1 had major cardio-myotoxic events in the three studied cohorts, inpatient monitoring may be preferred for a score of 1 or higher. Patients with score equal or above 2 may benefit most from empiric immunosuppression due to their >30% event rate. Further studies of this risk score may allow much-needed decision support for clinicians managing this rare and complex disease entity.

This study’s limitations should be recognized. The discovery cohort was derived from a multicenter registry wherein treating physicians adjudicated clinical data and outcomes. We attempted to mitigate this variability by excluding cases with no evidence for myocarditis per Bonaca’s criteria and by utilizing a core lab for ECG interpretation. Second, this cohort had missing data (detailed in Table 1) which were imputed. Imputation was necessary to power this analysis and prevented bias linked to potential non-random missingness. Finally, while the model tested known predictor variables, it is possible that unknown confounders were not included. Nevertheless, this risk score performed well in two independent external cohorts suggesting the validity of our model despite the aforementioned limitations.

In conclusion, we used a large multicenter registry to study predictors of severe outcomes in ICI-myocarditis and establish a risk score. This risk score was validated in 2 independent cohorts. These contemporary cohorts continue to show a high frequency of severe cardio-myotoxicity, occurring in one third of patients over the first month. A multivariable model showed that magnitude of troponin increase, pre-existing thymoma, presence of symptoms, low LVEF, and low QRS-voltage were associated with severe cardio-myotoxicity. These findings were used to derive a practical risk score which performed well in predicting adverse events in patients who present with ICI-myocarditis. This risk score was applied prospectively and was promising in identifying patients who could be safely managed without corticosteroids or other immunosuppressants.

## Data Availability

The international ICI myocarditis registry is open to the entire scientific community based on a 'reciprocity principle', i.e., access to the cohort will only be allowed to those entities also willing to share data. Access to the database is granted to researchers and clinicians who are part of the consortium and have been approved by the project's coordination (Pr Joe-Elie SALEM and Dr Javid MOSLEHI).

## Acknowledgements

none

## Author Information Statement

International ICI-Myocarditis Registry: Osnat Itzakhi Ben Zadok (USA), Anita Deswal (USA), Franck Thuny (France), Anissa Bouali (France), Andrew Hughes (USA), Lisa Moser (USA), Maxime Faure (France), Rocio Baro Vila (Canada), Jesus Jimenez (USA), Seyed Ebrahim Kassaian (USA), Elise Paven (France), Elena Galli (France), Teresa Lopez Fernandez (Spain); Lucia Cobarro (Spain), Manhal Habib (Israel), Kazuko Tajiri (Japan), Fanny Rocher (France), Theodora Bejan-Angoulvant (France), Mehmet Asim Bilen (USA), Susmita Parashar (USA), Avirup Guha (USA), Wenjing Song (China), David Koenig (Switzerland), Kirsten Mertz (Switzerland), Carrie Lenneman (USA), Andrew Haydon (Australia), Chloe Lesiuk (France), Romain Tresorier (France), Yazeed Samara (USA), Christian Grohe (Germany), Pierre Yves Dietrich (Switzerland), Sean Tierney (USA), Elie Rassy (Lebanon), Elvire Mervoyer (France), Shigeaki Suzuki (Japan), Satoshi Fukushima (Japan), Avirup Guha (USA), Maxime Robert-Halabi (Canada), Ryota Morimoto (Japan), Kazuko Tajiri (Japan), Robert Copeland-Halperin (USA), Michael Layoun (USA), Jun Wang (China), Suran Fernando (Australia), Eugenia Rota (Italy), Yumi Katsume (Japan), Yukiko Kiniwa (Japan), Ellen Warner (Canada), Nobuhiko Seki (Japan), Theresa Ruf (Germany), Jess DeLaune (USA), Nazanin Aghel (Canada)

## Supplementary

**Figure S1:**
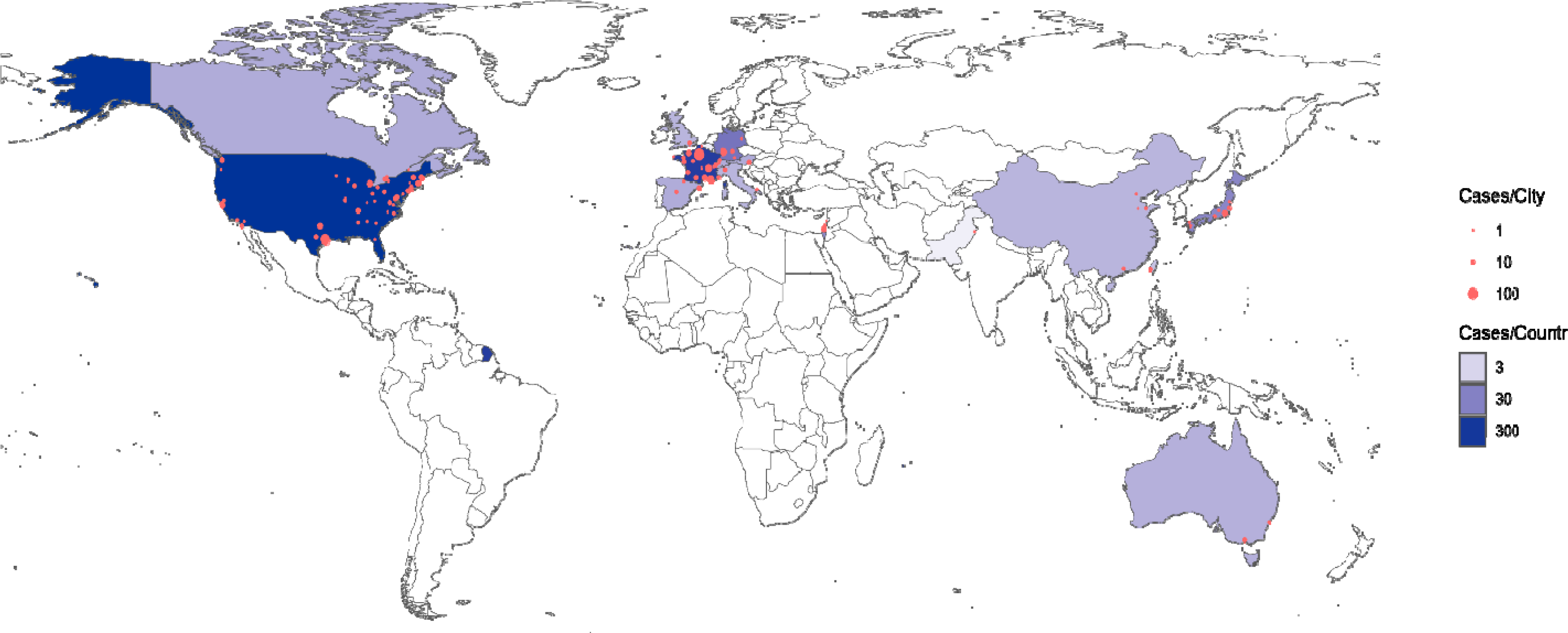
Geographic distribution of study population (discovery cohort)

**Figure S2:**
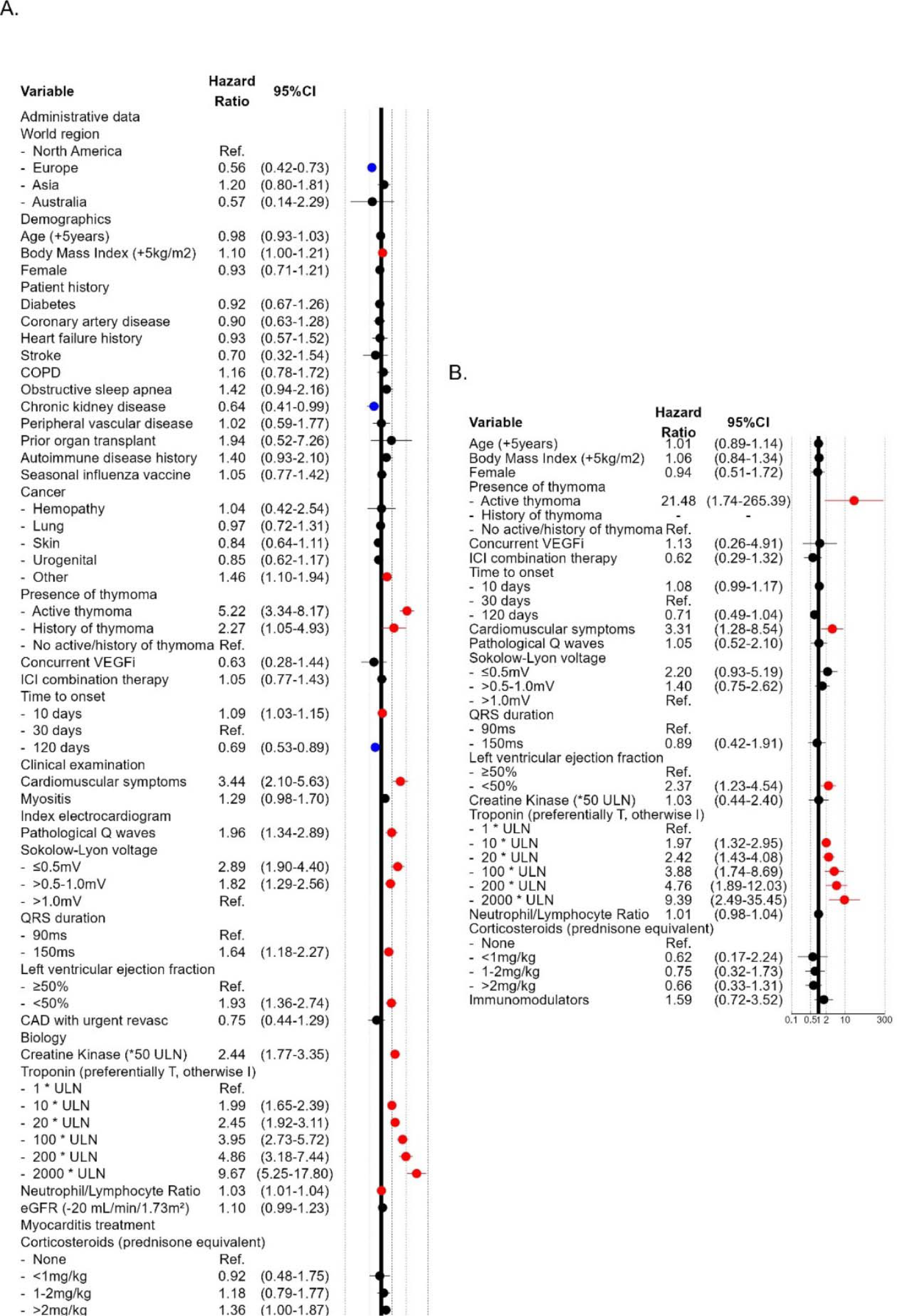
Univariate (A) and Unimputed (B) (n=238) multivariable models for association with major cardio-myotoxic events. *<u>Abbreviations</u>*: CAD: coronary artery disease, CI: confidence interval, COPD: Chronic obstructive pulmonary disease, eGFR: estimated glomerular filtration rate, ICI: immune checkpoint inhibitor, ms: millisecond, mV: millivolt, Ref: reference, ULN: upper limit of normal, VEGFi: Vascular Endothelial Growth Factor Inhibitor, Immunomodulator: non-steroidal immunomodulators only

**Figure S3:**
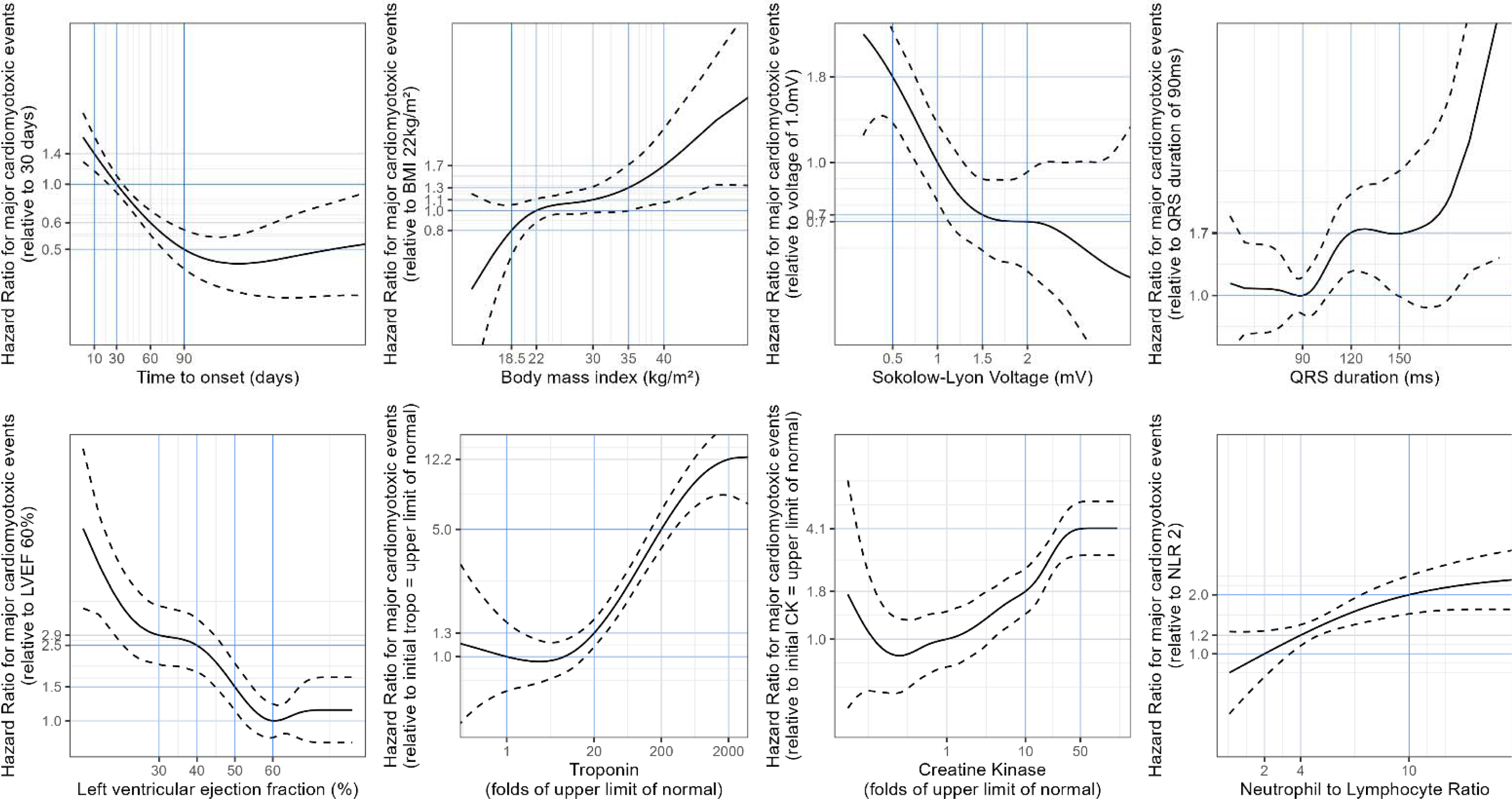
Spline models. Hazard-ratio for major cardio-myotoxic event at 30 days after presentation as a function of continuous variable predictors using data available at initial presentation. Troponin and creatine kinase were log transformed to account for non-linearity. *<u>Abbreviations:</u>* BMI: body mass index, LVEF: left ventricular ejection fraction, CK: creatine kinase, NLR: neutrophil to lymphocyte ratio

**Figure S4:**
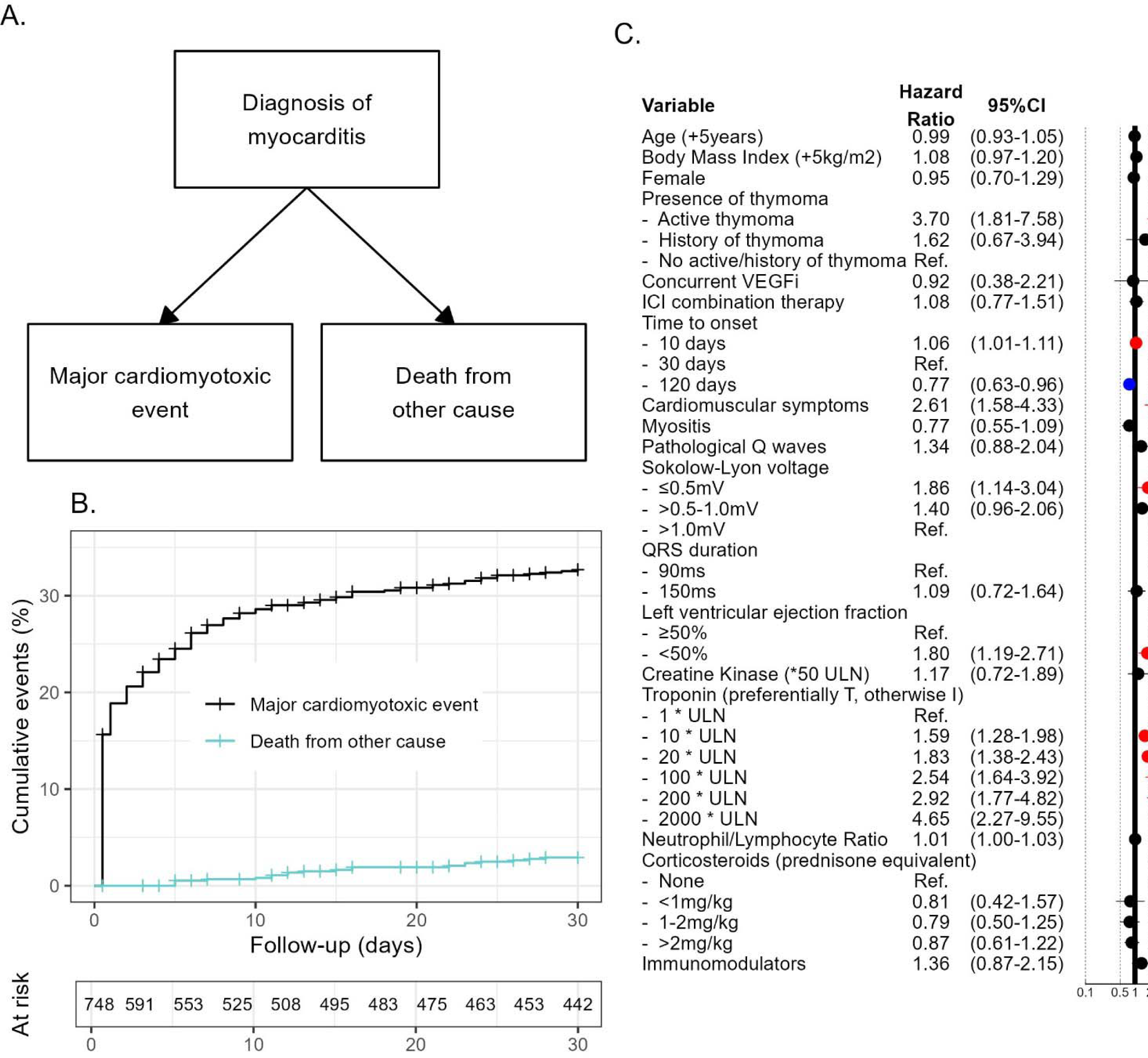
Predictors of 30-days mortality with competing risk approach. Illustration of competing risks for study patients (A). Cumulative incidence of competing risks during study period (B). Explanatory multivariable model for association with major cardio-myotoxic events fitted using Aalen Johanssen approach to account for death from other causes (C). *<u>Abbreviations</u>*: eGFR: estimated glomerular filtration rate, ICI: immune checkpoint inhibitor, ms: millisecond, mV: millivolt, Ref: reference, ULN: upper limit of normal, VEGFi: Vascular Endothelial Growth Factor Inhibitor, Immunomodulator: non-steroidal immunomodulators only

**Figure S5:**
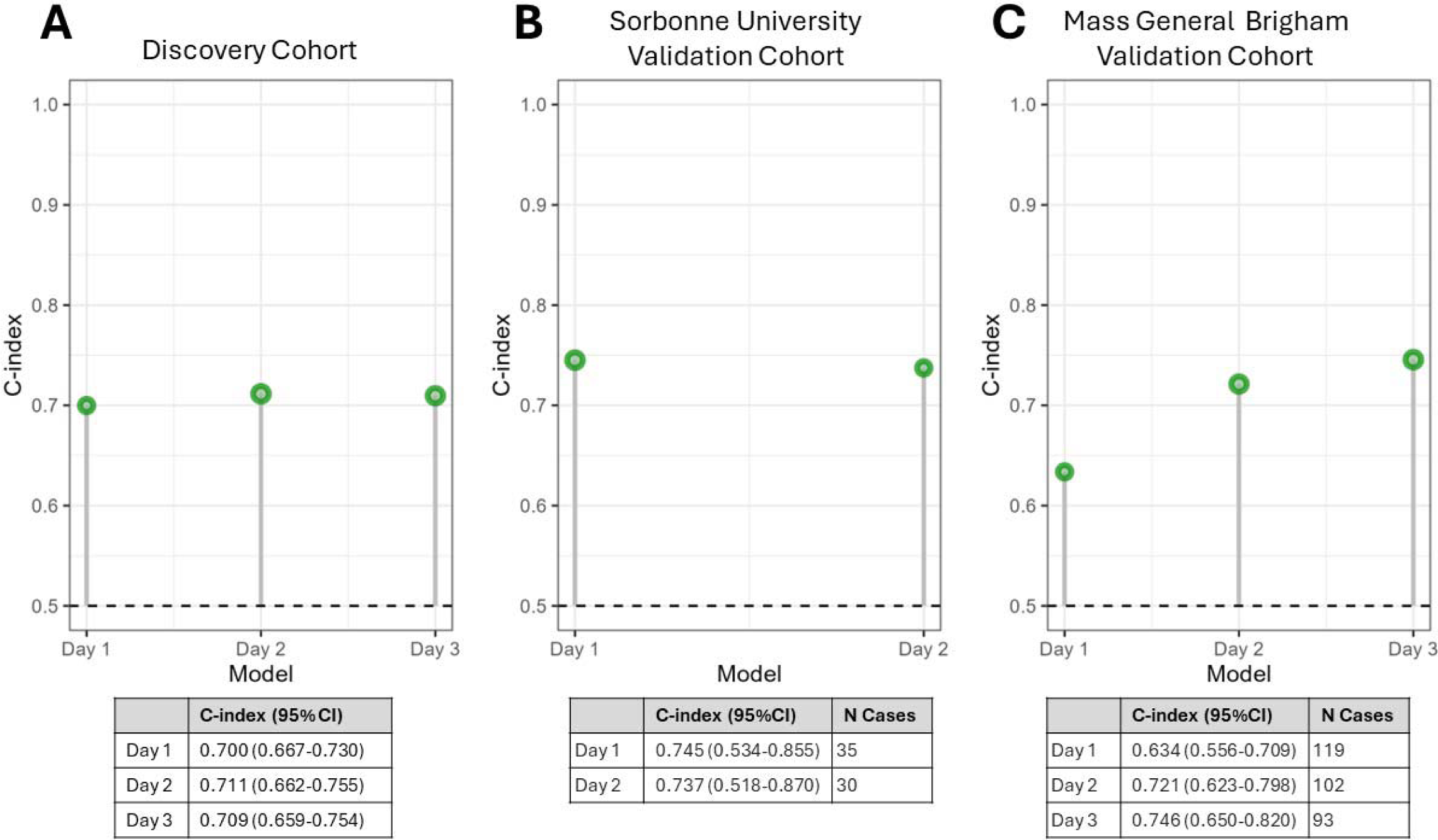
Harrel’s c-index of risk score for association with major cardio-myotoxic events. Graphical and tabular display of c-index when applied on a daily basis to Discovery Cohort (A), Sorbonne University Validation Cohort (B), Mass General Brigham Validation Cohort (C) *<u>Abbreviations:</u>* CI: confidence interval

**Figure S6:**
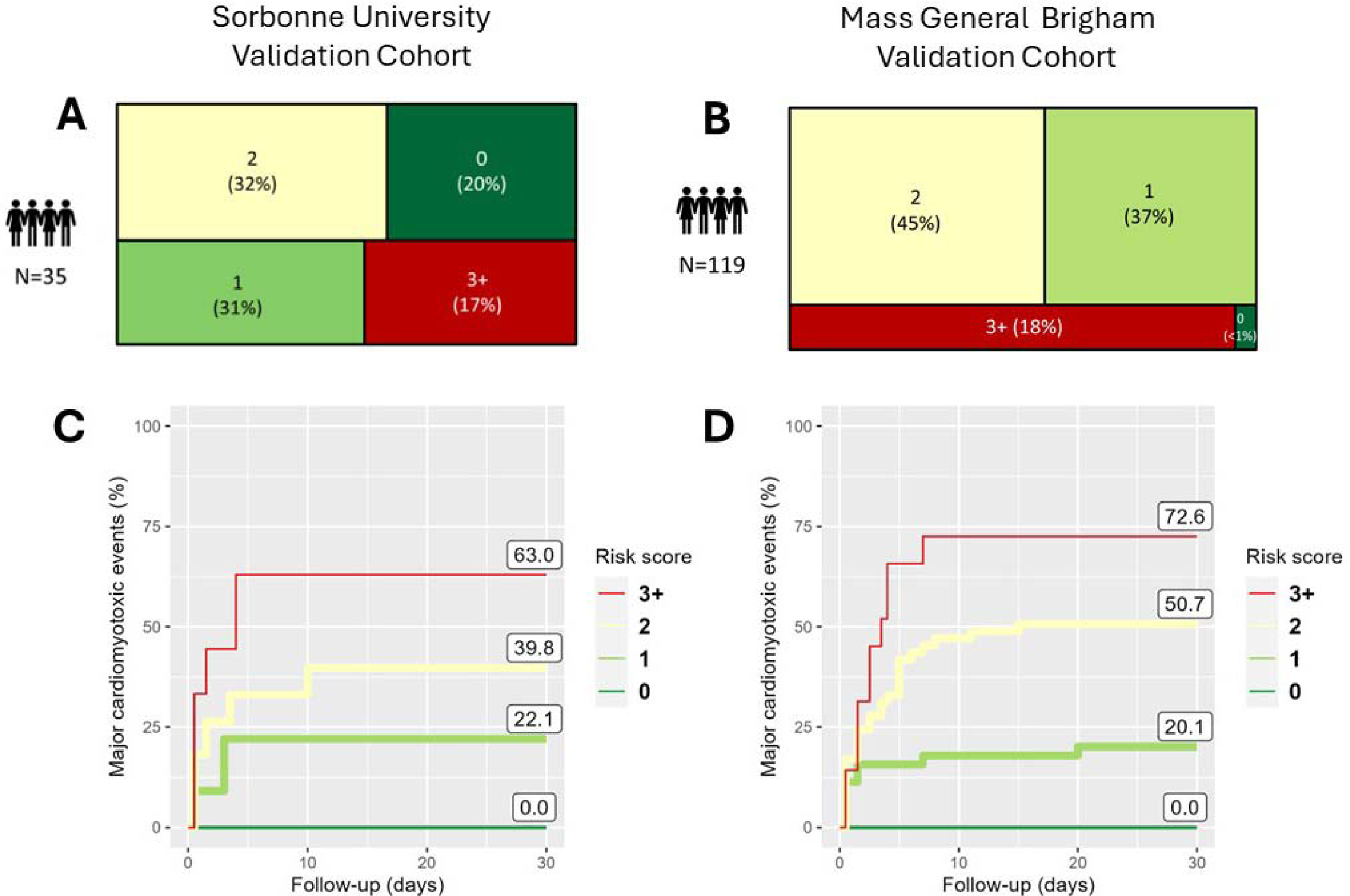
Validation cohorts risk score distribution and cardio-myotoxic event rate. Risk score distribution for Sorbonne University (A) and for Mass General Brigham (B) validation cohorts. Cumulative incidence of major cardio-myotoxic events according to for risk score level in the Sorbonne University (C) and in the Mass General Brigham Hospitals (D) validation cohorts

**Table S1:** Collaborators by institution (n=757 cases for discovery cohort, n=127 institutions, n=17 countries; and n=149 cases for replication cohorts, n=2 institutions, n=2 countries)

| Center | Country | City | Admin | N,<br>discovery<br>cohort | N,<br>validation<br>cohort | Collaborators |
| --- | --- | --- | --- | --- | --- | --- |
| Sorbonne University | France | Paris |  | 101 | 35 | Joe Elie Salem, Stephane Ederhy, Sonali Rao, Yves Allenbach, Thomas Similowski |
| Mass General Brigham | United States | Boston | Massachusetts | 1 | 119 | Anju Nohria, Osnat Ithzaki Ben Zadok |
| University of Texas MD Anderson Cancer Center | United States | Houston | Texas | 93 | 0 | Nicolas Palaskas, Anita Deswal |
| Heidelberg University Hospital | Germany | Heidelberg |  | 36 | 0 | Lorenz Lehmann |
| Aix-Marseille University, University Mediterranean Center of Cardio-Oncology | France | Marseille |  | 31 | 0 | Jennifer Cautela, Franck Thuny |
| Hospices Civils de Lyon | France | Lyon |  | 30 | 0 | Pierre-Yves Courand, Anissa Bouali |
| University of Michigan | United States | Ann Arbor | Michigan | 30 | 0 | Salim S. Hayek |
| Stanford University | United States | Stanford | California | 19 | 0 | Han Zhu |
| University of Texas Southwestern Medical Center | United States | Dallas | Texas | 16 | 0 | Vlad G. Zaha |
| University of Washington | United States | Seattle | Washington | 16 | 0 | Richard K. Cheng |
| Normandie Univ, UNICAEN, INSERM U1086 ANTICIPE; Caen-Normandy University Hospital. | France | Caen |  | 14 | 0 | Joachim Alexandre |
| CHU Montpellier | France | Montpellier |  | 14 | 0 | François Roubille |
| Yale School of Medicine | United States | New Haven | Connecticut | 14 | 0 | Lauren A. Baldassarre |
| University of California San Francisco | United States | San Francisco | California | 13 | 0 | Javid Moslehi, Alan H. Baik |
| Tel Aviv Sourasky Medical Center, School of Medicine, Tel Aviv University | Israel | Tel Aviv-Yafo |  | 12 | 0 | Michal Laufer-Peri |
| Beth Israel Deaconess Medical Center | United States | Boston | Massachusetts | 11 | 0 | Aarti Asnani |
| Maine Medical Center | United States | Portland | Maine | 10 | 0 | Sanjeev Francis |
| University of Virginia | United States | Charlottesville | Virginia | 10 | 0 | Elizabeth M. Gaughan |
| Vanderbilt University Medical Center | United States | Nashville | Tennessee | 10 | 0 | Douglas Johnson, Andrew M Hughes |
| APHP Hopital Lariboisiere | France | Paris |  | 9 | 0 | Guillaume Bailly |
| Dartmouth Hitchcock Medical Center | United States | Lebanon | New Hampshire | 9 | 0 | Danette L Flint |
| IUHW Mita Hospital | Japan | Tokyo |  | 9 | 0 | Yuichi Tamura |
| Medical University of Graz; BioTechMed Graz; St. Johann in Tirol General Hospital | Austria | Graz, St. Johann in Tirol |  | 9 | 0 | Peter Rainer, Lisa Moser |
| APHP Hopital Bichat | France | Paris |  | 8 | 0 | Dimitri Arangalage |
| CHU Rangueil Toulouse | France | Toulouse |  | 8 | 0 | Eve Cariou |
| Rabin Medical Center | Israel | Petah Tiqwa |  | 8 | 0 | Osnat Ithzaki Ben Zadok |
| University of California San Diego Health | United States | San Diego | California | 8 | 0 | Anna Narezkina, John Power |
| University of Utah | United States | Salt Lake | Utah | 7 | 0 | Roberta Florido |
|  |  | City |  |  |  |  |
| University of Texas at Austin<br>Ascension Seton | United States | Austin | Texas | 7 | 0 | Yan Liu |
| Barts Health NHS Trust | United Kingdom | London |  | 6 | 0 | Shanthini M Crusz |
| Maisonneuve-Rosemont Hospital | France | Bordeaux |  | 6 | 0 | Nahema Issa, Maxime Faure |
| CHU Nantes | France | Nantes |  | 6 | 0 | Nicolas Piriou |
| Mc Master University | Canada | Hamilton | Ontario | 6 | 0 | Darryl Leong, Rocio Baro Vila |
| Peter MacCallum Cancer Centre | Australia | Melbourne |  | 6 | 0 | Shahneen Sandhu |
| Disease Unit for Myocarditis and<br>Arrhythmogenic Cardiomyopathies,<br>IRCCS San Raffaele Scientific Institute | Italy | Milan |  | 6 | 0 | Giovanni Peretto |
| Washington University | United States | St. Louis | Missouri | 6 | 0 | Isik Turker, Jesus Jimenez |
| Hôpital Européen Georges Pompidou,<br>Université Paris Cité | France | Paris |  | 5 | 0 | Assié Eslami |
| APHP Hopital Henri Mondor | France | Créteil |  | 5 | 0 | Charlotte Fenioux |
| Bellvitge University Hospital – Catalan<br>Institute of Oncology, IDIBELL,<br>CIBERCV | Spain | Barcelona |  | 5 | 0 | Pedro Moliner |
| Centre Hospitalier Universitaire<br>Vaudois | Switzerland | Lausanne |  | 5 | 0 | Michel Obeid |
| Chi Mei Medical Center | Taiwan | Tainan |  | 5 | 0 | Wei Ting Chang |
| Northwestern University | United States | Chicago | Illinois | 5 | 0 | Nausheen Akhter |
| University of Wisconsin School of<br>Medicine and Public Health | United States | Madison | Wisconsin | 5 | 0 | Stephen M. Ewer |
| Medstar Washington Hospital Center | United States | Washington | District of Columbia | 4 | 0 | Sayed Ebrahim Kassaian |
| University Hospital Erlangen | Germany | Erlangen |  | 4 | 0 | Lucie Heinzerling |
| University of Pittsburgh Medical<br>Center | United States | Pittsburgh | Pennsylvania | 4 | 0 | Joshua E Levenson, Benay Ozbay |
| CHU Rennes | France | Rennes |  | 3 | 0 | Elise Paven, Elena Galli |
| Guangdong Academy of Medical<br>Sciences | China | Guangzhou |  | 3 | 0 |  |
| Hospital Universitario La Paz | Spain | Madrid |  | 3 | 0 | Teresa Lopez Fernandez, Lucia Cobarro |
| Rambam Medical Center | Israel | Haifa |  | 3 | 0 | Manhal Habib |
| Rosewell Park Cancer Center | United States | Buffalo | New York | 3 | 0 |  |
| University of Tsukuba | Japan | Kitaibaraki |  | 3 | 0 | Kazuko Tajiri |
| Center for Cancer Research National<br>Cancer Institute National Institutes of<br>Health | United States | Bethesda | Maryland | 2 | 0 |  |
| CHU Nice | France | Nice |  | 2 | 0 | Fanny Rocher |
| CHU Tours, University of Tours | France | Tours |  | 2 | 0 | Theodora Bejan-Angoulvant |
| Cleveland Clinic Foundation | United States | Cleveland | Ohio | 2 | 0 |  |
| Emory University | United States | Atlanta | Georgia | 2 | 0 | Mehmet Asim Bilen, Susmita Parashar |
| Houston Methodist Hospital | United States | Houston | Texas | 2 | 0 |  |
| Ohio State University Wexner Medical<br>Center | United States | Columbus | Ohio | 2 | 0 | Avirup Guha |
| The First Affiliated Hospital of Weifang<br>Medical University | China | Weifang |  | 2 | 0 | Wenjing Song |
| University Hospital Basel | Switzerland | Basel |  | 2 | 0 | David Koenig, Kirsten Mertz |
| University of Alabama Birmingham | United States | Birmingham | Alabama | 2 | 0 | Carrie Lenneman |
| University of California at Davis | United States | Davis | California | 2 | 0 |  |
| Vidant Medical Center/East Carolina University | United States | Greenville | North Carolina | 2 | 0 |  |
| Alfred Health, Monash University | Australia | Melbourne |  | 1 | 0 | Andrew Haydon |
| Allama Iqbal Medical College | Pakistan | Lahore |  | 1 | 0 |  |
| Anjo Kosei Hospital | Japan | Anjomachi |  | 1 | 0 |  |
| Baylor College of Medicine | United States | Houston | Texas | 1 | 0 |  |
| Bern University Hospital | Switzerland | Bern |  | 1 | 0 |  |
| Cedars-Sinai Medical Center Network | United States | Los Angeles | California | 1 | 0 | Anja Karlstaedt |
| Centre Léon Bérard | France | Lyon |  | 1 | 0 | Chloe Lesiuk |
| Charite Campus Mitte (CMM) | Germany | Berlin |  | 1 | 0 |  |
| CHU Brest | France | Brest |  | 1 | 0 |  |
| CHU Clermont Ferrand | France | Clermont-Ferrand |  | 1 | 0 | Romain Tresorier |
| CHU Lille | France | Lille |  | 1 | 0 |  |
| Clínica Universidad de Navarra | Spain | Pamplona |  | 1 | 0 |  |
| Cliniques universitaires Saint-Luc | Belgium | Bruxelles |  | 1 | 0 |  |
| Cooper University Hospital | United States | Camden | New Jersey | 1 | 0 |  |
| Dana Farber Cancer Institute | United States | Boston | Massachusetts | 1 | 0 |  |
| Eisenhower Medical Center | United States | Rancho Mirage | California | 1 | 0 | Yazeed Samara |
| ELK Thorax Center | Germany | Berlin |  | 1 | 0 | Christian Grohe |
| General Hospital of Chinese People's Liberation Army | China | Beijing |  | 1 | 0 |  |
| Geneva University Hospitals and Medical Faculty | Switzerland | Geneva |  | 1 | 0 | Pierre Yves Dietrich |
| Hartford Hospital | United States | Hartford | Connecticut | 1 | 0 |  |
| Heart Care Centers of Illinois | United States | Chicago | Illinois | 1 | 0 | Sean Tierney |
| Hôtel Dieu de France University Hospital | Lebanon | Beirut |  | 1 | 0 | Elie Rassy |
| ICO Nantes | France | Nantes |  | 1 | 0 | Elvire Mervoyer |
| Institut Bergonié Centre de Lutte contre le Cancer | France | Bordeaux |  | 1 | 0 |  |
| Japan Community Health Organization Kyushu Hospital | Japan | Fukuoka |  | 1 | 0 |  |
| Keio University | Japan | Tokyo |  | 1 | 0 | Shigeaki Suzuki |
| Kumamoto University | Japan | Kumamoto |  | 1 | 0 | Satoshi Fukushima |
| Lahey Hospital and Medical Center | United States | Burlington | Massachusetts | 1 | 0 |  |
| Markey Cancer Center, University of Kentucky | United States | Lexington | Kentucky | 1 | 0 |  |
| Marshall University | United States | Huntington | West Virginia | 1 | 0 |  |
| Mayo Clinic | United States | Rochester | Minnesota | 1 | 0 |  |
| Georgia Cancer Center, Augusta University | United States | Augusta | Georgia | 1 | 0 | Avirup Guha |
| Memorial Sloan Kettering Weill Cornell Medical College | United States | New York | New York | 1 | 0 |  |
| Mitsui Memorial Hospital | Japan | Tokyo |  | 1 | 0 |  |
| Montreal Heart Institute | Canada | Montreal | Quebec | 1 | 0 | Maxime Robert-Halabi |
| Mount Vernon Cancer Centre | United Kingdom | London |  | 1 | 0 |  |
| Nagoya University Graduate School of Medicine | Japan | Nagoya |  | 1 | 0 | Ryota Morimoto |
| National Cancer Center Hospital East | Japan | Kashiwa |  | 1 | 0 | Kazuko Tajiri |
| New York Institute of Technology College of Osteopathic Medicine | United States | New York | New York | 1 | 0 |  |
| New York-Presbyterian Brooklyn Methodist Hospital | United States | New York | New York | 1 | 0 |  |
| Northwell Health | United States | New York | New York | 1 | 0 | Robert Copeland-Halperin |
| Providence St. Joseph Health | United States | Portland | Oregon | 1 | 0 | Michael Layoun |
| Qianfoshan Hospital | China | Jinan |  | 1 | 0 | Jun Wang |
| Royal North Shore Hospital | Australia | Sydney |  | 1 | 0 | Suran Fernando |
| Saiseikai Narashino Hospital | Japan | Narashino |  | 1 | 0 |  |
| San Giacomo Hospital | Italy | Monopoli |  | 1 | 0 | Eugenia Rota |
| Sendai Kousei Hospital | Japan | Sendai |  | 1 | 0 | Yumi Katsume |
| Shinshu University | Japan | Matsumoto |  | 1 | 0 | Yukiko Kiniwa |
| St. Luke's International Hospital | Japan | Tokyo |  | 1 | 0 |  |
| Sunnybrook Odette Cancer Centre | Canada | Toronto |  | 1 | 0 | Ellen Warner |
| Teikyo University School of Medicine | Japan | Tokyo |  | 1 | 0 | Nobuhiko Seki |
| Tokyo to Saiseikai Central Hospital | Japan | Tokyo |  | 1 | 0 |  |
| Tokyo Women's Medical University | Japan | Tokyo |  | 1 | 0 |  |
| University Hospital LMU | Germany | Munich |  | 1 | 0 | Lucie Heinzerling, Theresa Ruf |
| University Hospital Mainz | Germany | Mainz |  | 1 | 0 |  |
| University of Chicago | United States | Chicago | Illinois | 1 | 0 |  |
| University of Florida | United States | Gainesville | Florida | 1 | 0 | Jess DeLaune |
| University of Massachusetts, Berkshire Medical Centre | United States | Pittsfield | Massachusetts | 1 | 0 |  |
| University of North Carolina Medical Center Chapel Hill | United States | Chapel Hill | North Carolina | 1 | 0 |  |
| University of southern California | United States | Los Angeles | California | 1 | 0 |  |
| University of Toronto | Canada | Toronto |  | 1 | 0 | Nazanin Aghel |
| VCU Medical center | United States | Richmond | Virginia | 1 | 0 |  |
| Veterans Affairs Puget Sound Health Care System-Seattle | United States | Seattle | Washington | 1 | 0 |  |

**Table S2:** Clinical Characteristics of Validation Cohorts.

|  | Sorbonne University, Paris (France) | Mass General Brigham, Boston (USA) |
| --- | --- | --- |
| **Presentation date periods** | April–2023 - April–2024 | January–2015 - June 2022 |
| **Sample size** | n=35, prospective | n=119, retrospective<sup>19</sup> |
| **Age (years, median, (IQR))** | 74 (71 - 79) | 73 (67 - 76) |
| **Female (n/N) (%)** | 15 (43%) | 40 (34%) |
| **ICI Regimen** |  |  |
| Anti-PD1 or anti-PDL1 monotherapies | 33 (94%) | 90 (76%) |
| Anti-CTLA4 monotherapy | 0 (0%) | 3 (3%) |
| Anti-CTLA4 & anti-PD1/PDL1 combination therapy | 2 (6%) | 26 (22%) |
| **Cancer type** |  |  |
| Hemopathy | 1 (3%) | 2 (2%) |
| Lung | 8 (23%) | 30 (25%) |
| Skin | 13 (37%) | 36 (30%) |
| Urogenital | 9 (26%) | 28 (24%) |
| Other cancers | 4 (11%) | 23 (19%) |
| **Time from 1<sup>st</sup> ICI infusion to myocarditis (days, median, (IRQ))** | 42 (27-79) | 52 (26 - 120) |
| **Major Cardiomyotoxic Events (30-days)** | 11 (31%) | 49 (41%) |
| Respiratory muscle failure requiring ventilation | 7 (20%) | 6 (5%) |
| Cardiomyotoxic death | 0 (0%) | 10 (8%) |
| Heart Failure requiring intravenous drugs | 4 (11%) | 30 (25%) |
| Severe arrhythmia* | 5 (14%) | 18 (15%) |
| All-cause mortality (30-days) | 0 (0%) | 18 (15%) |
| **Risk score parameters within the three first days (day 1 to 3)** |  |  |
| Active thymoma | 1 (3%) | 0 (0%) |
| Cardiomuscular symptoms | 24 (69%) | 117 (98%) |
| Sokolow Lyon index $\leq 0.5$ mV | 3 (9%) | 9 (8%) |
| LVEF<50% | 6 (17%) | 41 (34%) |
| Maximum Troponin** (fold upper limit of normal), median (IQR) | 26 (10 - 86) | 13 (4 - 84) |
| **Diagnostic certainty per updated Bonaca's criteria<sup>5</sup>** |  |  |
| Definite myocarditis | 35 (100%) | 37 (31%) |
| Probable myocarditis | 0 (0%) | 31 (26%) |
| Possible myocarditis | 0 (0%) | 51 (43%) |

\* Severe arrhythmia: defined as sustained ventricular arrhythmia, complete heart block, sudden cardiac death, use of atropine or isoproterenol, and/or use of pacemaker/defibrillator
\*\* preferentially cardiac troponin-T, if unavailable cardiac troponin I

**Table S3:** Diagnostic properties of the risk score to predict major cardio-myotoxic events at 30 days.

| Risk Score | Sensitivity | Specificity | Positive Predictive Value | Negative Predictive Value |
| --- | --- | --- | --- | --- |
| Discovery Cohort: International Redcap |  |  |  |  |
| 0+ | 1.00 (0.98-1.00) | 0.00 (0.00-0.01) | 0.32 (0.29-0.36) |  |
| 1+ | 0.99 (0.97-1.00) | 0.14 (0.11-0.17) | 0.35 (0.32-0.39) | 0.97 (0.90-1.00) |
| 2+ | 0.82 (0.77-0.87) | 0.48 (0.44-0.52) | 0.43 (0.39-0.48) | 0.85 (0.80-0.89) |
| 3+ | 0.46 (0.40-0.53) | 0.84 (0.81-0.87) | 0.59 (0.52-0.65) | 0.77 (0.73-0.80) |
| 4+ | 0.18 (0.14-0.24) | 0.98 (0.96-0.99) | 0.80 (0.68-0.89) | 0.71 (0.68-0.75) |
| 5+ | 0.06 (0.03-0.09) | 1.00 (0.99-1.00) | 0.91 (0.65-0.99) | 0.69 (0.65-0.72) |
| 6+ | 0.01 (0.00-0.04) | 1.00 (0.99-1.00) | 0.96 (0.35-0.99) | 0.68 (0.64-0.71) |
| Validation Cohort: Sorbonne University |  |  |  |  |
| 0 | 1.00 (0.70-1.00) | 0.00 (0.00-0.16) | 0.31 (0.18-0.48) |  |
| 1 | 1.00 (0.70-1.00) | 0.29 (0.15-0.49) | 0.39 (0.24-0.58) | 1.00 (0.60-1.00) |
| 2 | 0.82 (0.51-0.96) | 0.67 (0.47-0.82) | 0.53 (0.31-0.74) | 0.89 (0.66-0.98) |
| 3 | 0.36 (0.15-0.65) | 0.92 (0.73-0.99) | 0.67 (0.30-0.91) | 0.76 (0.58-0.88) |
| 4 | 0.18 (0.04-0.49) | 0.96 (0.78-1.00) | 0.67 (0.20-0.94) | 0.72 (0.54-0.85) |
| 5 | 0.00 (0.00-0.30) | 1.00 (0.84-1.00) |  | 0.69 (0.52-0.82) |
| 6 | 0.00 (0.00-0.30) | 1.00 (0.84-1.00) |  | 0.69 (0.52-0.82) |
| Validation Cohort: Mass General Brigham |  |  |  |  |
| 0 | 1.00 (0.91-1.00) | 0.00 (0.00-0.06) | 0.41 (0.33-0.50) |  |
| 1 | 1.00 (0.91-1.00) | 0.01 (0.00-0.08) | 0.42 (0.33-0.51) | 1.00 (0.17-1.00) |
| 2 | 0.80 (0.66-0.89) | 0.50 (0.39-0.61) | 0.53 (0.41-0.64) | 0.78 (0.64-0.88) |
| 3 | 0.29 (0.18-0.43) | 0.90 (0.80-0.95) | 0.67 (0.45-0.83) | 0.64 (0.54-0.73) |
| 4 | 0.02 (0.00-0.12) | 0.99 (0.92-1.00) | 0.50 (0.09-0.91) | 0.59 (0.50-0.67) |
| 5 | 0.00 (0.00-0.09) | 1.00 (0.94-1.00) |  | 0.59 (0.50-0.67) |
| 6 | 0.00 (0.00-0.09) | 1.00 (0.94-1.00) |  | 0.59 (0.50-0.67) |

